# Mapping evidence on cryptococcal antigen infection among HIV-infected persons in sub-Saharan Africa- A Scoping Review Protocol

**DOI:** 10.1101/2023.02.03.23285416

**Authors:** Khululiwe Dlamini, Boitumelo Moetlhoa, Astrid Turner, Kuhlula Maluleke, Tivani Mashamba-Thompson

## Abstract

**Introduction:** Infections of the central nervous system are a considerable basis of mortality in people living with Human immunodeficiency virus (HIV), with progression to cryptococcal meningitis documented at around 15% of HIV-associated mortality globally, with nearly three-quarters occurring in the sub-Saharan Africa. Discoveries from previous studies preluded that mortality amid cryptococcal antigen (CrAg) positive persisted to be elevated than in CrAg negative persons. One feasible interpretation of this could be due to undiagnosed cryptococcus. Laboratory investigations towards prompt identification of cryptococcal disease prior to cryptococcal meningitis has progressed to point-of-care testing with high sensitivity and specificity as seen with the CrAg lateral flow assay screening to expedite treatment. The aim of the study is to map and translate evidence on CrAg infection among HIV-infected persons in sub-Saharan Africa (SSA).

**Methodology:** The proposed scoping review will be conducted using guidelines proposed by Arksey and O’Malley methodological framework and Levac et al advanced method. It will be guided by the Preferred Reporting Items for Systematic Reviews and Meta-Analysis for Scoping Reviews. A comprehensive literature search of studies published from the first relevant publication to 2022 will be conducted on multiple electronic databases. Additional sources (grey literature) will also be searched. The search strategy will be generated and implemented by the principal investigator with assistance from a subject specialist, and an information specialist. Two reviewers will screen eligible studies. The screening will be guided by an inclusion and exclusion criteria. The mixed method appraisal tool (MMAT) version 2018 will be used to appraise the quality of the empirical studies.

**Discussion:** The proposed scoping review will map and translate evidence on CrAg infection among HIV-infected persons in sub-Saharan Africa. Synthesising and sharing recent evidence in this area has potential to help guide future research and interventions aimed at improving the management of CrAg infection among HIV-infected persons in sub-Saharan Africa and other high HIV-burdened settings.

## Introduction

Cryptococcal disease is a paramount cause of illness in people living with HIV/AIDS and has major consequences if left undiagnosed and untreated. An approach to identifying risk of progression to disease is necessary to guide CrAg screening in the distribution of resources particularly in resource-limited settings when supervising people demonstrating advanced HIV disease. Updated guidelines are required to provide recommendations on approach to detection, diagnosis and treatment. These guidelines also include strategies on preventing invasive illness and potential impact with antiretroviral therapy (ART). When cryptococcal disease is suspected, the CrAg point-of-care (POC) testing is conducted to enable prompt treatment before the progression to cryptococcal meningitis (CM). CM is the most common appearance of the infection and a contributing factor to the morbidity and mortality in the SSA. Mortality is further increased through the immediate initiation of ART in people living with HIV (PLHIV) that have been diagnosed with CM. The guidelines recommended by the World Health Organisation (WHO), is to delay ART by 4–6 weeks from the initiation of antifungal treatment (1, 2).

Discovering the estimates of incidence and prevalence of HIV-related diseases specially in cryptococcal infection has been continuous in studies globally (3–6), despite the advancement of HIV management and medicine. The global burden estimates have important purpose in recommended guidelines for prevention strategies required for treatment. Studies were conducted on “the prevalence of cryptococcal infection among HIV-persons with a CD4 cell count of less than 100 cells/μL” however, the clinical characteristics of PLHIV, with the progression to CM proportional with elevated CD4 cell counts (101-200 cells/μL), would contribute discernment into the role of the immune response to the pathogenesis of CM. This enables customization of applicable treatment interventions for cryptococcosis (1). Identified patients begin pre-emptive therapy with a high concentrated dosage of fluconazole to inhibit development of severe disease (1, 7, 8). A CD4 cell count is a necessary tool in aiding decision-makers in clinical management (9). A CD4 cell count of less than 200 cells/μL is regarded a threshold for potential morbidity (10). The WHO recommends screening for CrAg in PLHIV, followed by pre-emptive antifungal treatment for a positive screening, to prevent progression to CM. This recommendation is for adults and adolescents living with HIV before the initiation or re-initiation of ART therapy (2). One study publicized that in Africa, the cryptococcosis-associated immune reconstitution inflammatory syndrome (IRIS) is dangerous with 27-83% reported mortalities, this would be due to undiagnosed cryptococcosis. Additional studies would be necessary to further understand the perseverance of high mortality in the context of establishing appropriate strategies in screening for CM within routine testing (7). Laboratory investigations towards early detection of cryptococcal disease prior to CM has progressed to POC testing with high sensitivity and specificity as seen with the CrAg lateral flow assay (CrAg LFA) screening to expedite treatment (9, 11), therefore the level of evidence to validate clinic-based CrAg LFA testing in sub-Saharan Africa (SSA) is not clear (12). The proposed scoping review aims to systematically map evidence on CrAg infection among HIV-infected persons in sub-Saharan Africa. It is anticipated that findings from this study will contribute evidence to propose improved management of CrAg infections within healthcare in SSA.

## Methodology

The scoping review will be performed using guidelines proposed by Arksey and O’Malley (14) methodological framework and Levac et al advanced method (15) and guided by the Preferred Reporting Items for Systematic Reviews and Meta-Analysis for Scoping Reviews (PRISMA-ScR) (16, 17). The five stages of Arksey and O’Malley framework that will be used to perform this scoping review are: i) identifying the research question ii) identifying relevant studies iii) eligible study selection iv) charting the data and v) collating, summarising and reporting the results. This scoping review will not incorporate stage six involving stakeholders’ consultation.

### i. Identifying the research question

The research question is: What is the level of evidence on CrAg infection among HIV-infected persons in SSA? To Refine the research question for the scoping review we will use the Population, Concept, Context (PCC) nomenclature summarized in table 1 below.

**Table 1:** PCC framework for refining the research question

|  |  |
| --- | --- |
| **Population** | HIV-infected persons<br>all infected people |
| **Concept** | CrAg infection<br>Opportunistic fungal agent resulting in HIV-related infection (18) |
| **Context** | sub-Saharan Africa |

### ii. Identifying relevant studies

Comprehensive literature searches of studies published from the first relevant publication to 2022 will be conducted on the following electronic databases from Scopus, PubMed, and EBSCO host (MEDLINE, Cumulative Index to Nursing and Allied Health Literature (CINAHL). Additionally, we will look at grey literature (university dissertations and thesis), government and international organisations such as WHO reports to determine more sources of information that have not been arranged by electronic databases. We will manually search for references cited in the included articles. The English language will be applied.

A comprehensive literature search strategy combining terms will be done in three separate searches: “HIV” “PLHIV” “HIV infected persons” “advance HIV disease” “HIV related disease”, “Cryptococcal” “Cryptococcus” “Cryptococcosis” “Cryptococcal antigen” “Cryptococcal antigen infection” “CrAg” “CrAg screening” “Cryptococcal meningitis” “Meningitis”, and “Sub-Saharan Africa” “SSA”.

The search strategy will be created and implemented by the principal investigator (PI) with assistance from a subject specialist, and an information specialist (university librarian). Each database search will be on record to show the following: date of search, electronic database, keywords/MeSH terms, and the number of recovered studies. A pilot search was conducted on one of the electronic databases and the findings are documented in table 2 below. To confirm accurate use of terminology, keywords may be edited to ensure database search suitability.

**Table 2:** Results of pilot search

| Date of search | Electronic Database | Keywords/MeSH terms | Number of hits |
| --- | --- | --- | --- |
| 05/10/2022 | PubMed | HIV: "hiv"[MeSH Terms] OR "hiv"[All Fields]<br>cryptococcal: "cryptococcus"[MeSH Terms] OR "cryptococcus"[All Fields] OR "cryptococcal"[All Fields]<br>infection: "infect"[All Fields] OR "infectability"[All Fields] OR "infectable"[All Fields] OR "infectant"[All Fields] OR "infectants"[All Fields] OR "infected"[All Fields] OR "infecteds"[All Fields] OR "infectibility"[All Fields] OR "infectible"[All Fields] OR "infecting"[All Fields] OR "infection's"[All Fields] OR "infections"[MeSH Terms] OR "infections"[All Fields] OR | 485 |
|  |  | "infection"[All Fields] OR<br>"infective"[All Fields] OR<br>"infectiveness"[All Fields] OR<br>"infectives"[All Fields] OR<br>"infectivities"[All Fields] OR<br>"infects"[All Fields] OR<br>"pathogenicity"[Subheading] OR<br>"pathogenicity"[All Fields] OR<br>"infectivity"[All Fields]<br>sub-Saharan Africa: "africa south of the sahara"[MeSH Terms] OR<br>("africa"[All Fields] AND "south"[All Fields] AND "sahara"[All Fields])<br>OR "africa south of the sahara"[All Fields] OR ("sub"[All Fields] AND "saharan"[All Fields] AND "africa"[All Fields]) OR "sub saharan africa"[All Fields] |  |

### iii. Eligible study selection

The following criteria will be used to select relevant studies:

#### Inclusion criteria

✓ Studies reporting evidence of HIV/AIDS patients
✓ Studies conducted among adults and children in SSA
✓ Studies reporting evidence on CM diagnosis from CSF sample and CrAg infection from whole blood, serum and plasma (CSF sample has the highest diagnostic value irrespective of the blood sample result)
✓ Studies reporting evidence on estimates of the incidence, prevalence, and mortality of cryptococcal infection and CM
✓ Studies reporting evidence on patients with historical cryptococcosis or overt clinical CM

#### Exclusion criteria

The following criteria will be used to exclude studies:

✓ Studies reporting evidence other than whole blood, CSF sample, serum and plasma
✓ Studies reporting evidence of patients receiving antifungal treatment
✓ Studies that report evidence of disease unrelated to CM (bacterial or viral meningitis, tuberculous meningitis, protozoan infections, and space-occupying cerebral oedema lesion e.g. tuberculoma, hydrocephalus or CNS malignancy)
✓ Studies reporting evidence on primary studies conducted outside SSA
✓ Studies that do not specify CM diagnosis, CrAg infection, or record of test performed
✓ Review articles

Following the electronic database search, the eligible studies will be transferred to Endnote, a citation management software. The PI will work independently to screen titles using the inclusion and exclusion criteria guide. The PI, with the assistance of an independent co-screener will screen the abstracts. Disagreements between the two screeners will be settled by a discussion until unanimity is reached. Screening of full article will be conducted by the same set of screeners. Any discrepancies that emerge after screening full articles will be settled by introducing a third screener. The degree of agreement amongst the screeners after full article screening will be established through calculating the Cohen’s Kappa Statistics (19, 20). The process of the study search decision will be guided by the PRISMA flow chart as demonstrated in figure 1 (16) to aid reporting of the scoping review.

### iv. Charting the data

A data charting form as outlined in table 3, will be used to derive information from each included study. Two independent reviewers will pilot test and modify the data charting form.

**Table 3:** Data charting form

|  |
| --- |
| **Author & Year of publication** |
| **Journal** |
| **Aim of study** |
| **Study population** |
| **Study setting (rural and urban)** |
| **Study location (sub-Saharan Africa)** |
| **Study design** |
| **Main findings** |
| **Other significant findings** |

### v. Collating, summarising and reporting the results

Findings will be presented in two ways. Firstly, from numerical data on the incidence, prevalence and mortality of the study population will be reported. Secondly, we will produce tables and graphs based from thematic analysis that originate from the PCC to guide a summary of themes to extract from findings in relation to answering the study question.

### vi. Appraisal of quality of evidence

The mixed method appraisal tool (MMAT) version 2018 will be utilized to appraise the quality of empirical studies, these are primary research acquired from observation, experimental or simulation of study methods i.e. quantitative, qualitative, and/or mixed method studies (21). Appraisal of the included studies will be intended to assess the objectives of the study, method, study design, candidate recruitment, data collected, analysis of data, discovered results, and authors’ discussions and conclusions to ensure they are reliable, trustworthy and valid. The appraisal process will involve two independent reviewers assessing the checklist and criterion based on methodology according to the MMAT (21). A percentage score will be calculated to evaluate the quality of studies selected as follows: i) ≤ 50%, will be low quality ii) 51–75%, will be average quality and iii) 76–100%, will be considered as high quality.

### vii. Ethical considerations

This scoping review involves synthesis of current evidence therefore ethical approval is not necessary. However, since the study is being conducted for degree purposes, ethical approval was sought. The study was ethically reviewed and approved submitted to the University of Pretoria Research Ethics Committee (Approval number: 606/2022).

## Discussion

Screening of the cryptococcal antigen is the preferred approach for identifying risk of progression to disease when managing people presenting with advanced HIV infection (2). Screening for CrAg in PLHIV, followed by pre-emptive antifungal treatment for a positive screening are one of the WHO’s recommendations for prevention of progression to CM (2). Highly accurate CrAg POC tests have been used for disease screening to help expedite treatment (9, 11).

This scoping review will exclude studies conducted outside SSA as they will not be reflecting findings associated with the region of interest. Studies reporting evidence of patients receiving antifungal treatment will also be excluded as they will be giving evidence on people who have already been screened and not on the CrAg infection among PLHIV. Furthermore, review articles will be excluded on literature from primary studies conducted by other researchers. We anticipate that results of this scoping review will guide future policy and practice aimed on improving the HIV management.

## Data Availability

No datasets were generated or analysed during the current study. All relevant data from this study will be made available upon study completion.

## Acknowledgement

The authors acknowledge the support received from the University of Pretoria during the development of this protocol. The authors would also like to thank the library services for their assistance with optimising the search strategy.

## Supporting material

S1 Figure 1: PRISMA-Scr flow chart

## References

1. Tugume L, Rhein J, Hullsiek KH, Mpoza E, Kiggundu R, Ssebambulidde K, et al HIV-Associated Cryptococcal Meningitis Occurring at Relatively Higher CD4 Counts. J Infect Dis. 2019;219(6):877–83.

2. World Health Organization. Guidelines for the diagnosis, prevention, and management of cryptococcal disease in HIV-infected adults, adolescents and children, March 2018: supplement to the 2016 consolidated guidelines of the use of antiretroviral drugs for treating and preventing HIV infection [Publications]. World Health Organization; 2018 [updated 2018. Available from: https://apps.who.int/iris/handle/10665/260399.

3. Longley N, Jarvis JN, Meintjes G, Boulle A, Cross A, Kelly N, et al Cryptococcal Antigen Screening in Patients Initiating ART in South Africa: A Prospective Cohort Study. Clin Infect Dis. 2016;62(5):581–7.

4. Li Y, Huang X, Chen H, Qin Y, Hou J, Li A, et al The prevalence of cryptococcal antigen (CrAg) and benefits of pre-emptive antifungal treatment among HIV-infected persons with CD4+ T-cell counts < 200 cells/μL: evidence based on a meta-analysis. BMC Infect Dis. 2020;20(1):410.

5. Ford N, Meintjes G, Vitoria M, Greene G, Chiller T. The evolving role of CD4 cell counts in HIV care. Curr Opin HIV AIDS. 2017;12(2):123–8.

6. Ford N, Shubber Z, Jarvis JN, Chiller T, Greene G, Migone C, et al CD4 Cell Count Threshold for Cryptococcal Antigen Screening of HIV-Infected Individuals: A Systematic Review and Metaanalysis. Clin Infect Dis. 2018;66(suppl_2):S152–s9.

7. Otto SBJ, George PE, Mercedes R, Nabukeera-Barungi N. Cryptococcal meningitis and immune reconstitution inflammatory syndrome in a pediatric patient with HIV after switching to second line antiretroviral therapy: a case report. BMC Infect Dis. 2020;20(1):68.

8. Rajasingham R, Wake RM, Beyene T, Katende A, Letang E, Boulware DR. Cryptococcal Meningitis Diagnostics and Screening in the Era of Point-of-Care Laboratory Testing. J Clin Microbiol. 2019;57(1).

9. Drain PK, Hong T, Krows M, Govere S, Thulare H, Wallis CL, et al Validation of clinic-based cryptococcal antigen lateral flow assay screening in HIV-infected adults in South Africa. Scientific Reports. 2019;9(1):2687.

10. Govender NP, Roy M, Mendes JF, Zulu TG, Chiller TM, Karstaedt AS. Evaluation of screening and treatment of cryptococcal antigenaemia among HIV-infected persons in Soweto, South Africa. HIV Med. 2015;16(8):468–76.

11. Chukwuanukwu RC, Uchenna N, Mbagwu SI, Chukwuanukwu TO, Charles O. Cryptococcus neoformans seropositivity and some haematological parameters in HIV seropositive subjects. Journal of Infection and Public Health. 2020;13(7):1042–6.

12. Rajasingham R, Smith RM, Park BJ, Jarvis JN, Govender NP, Chiller TM, et al Global burden of disease of HIV-associated cryptococcal meningitis: an updated analysis. Lancet Infect Dis. 2017;17(8):873–81.

13. National Institute for Communicable Disease. Cryptococcal antigen screening surveillance report, South Africa, February 2017-July 2019 - Google Search 2022 [Available from: https://www.google.com/search?q=Cryptococcal+antigen+screening+surveillance+report%2C+South+Africa%2C+February+2017+%E2%80%93+July+2019&rlz=1C1CAFB_enZA626ZA626&oq=Cryptococcal+antigen+screening+surveillance+report%2C+South+Africa%2C+February+2017+%E2%80%93+July+2019&aqs=chrome..69i57j69i59.1762438j0j4&sourceid=chrome&ie=UTF-8.

14. Arksey H, O’Malley L. Scoping studies: towards a methodological framework. International Journal of Social Research Methodology. 2005;8(1):19–32.

15. Levac D, Colquhoun H, O’Brien KK. Scoping studies: advancing the methodology. Implement Sci. 2010;5(1):1–9.

16. Page MJ, McKenzie JE, Bossuyt PM, Boutron I, Hoffmann TC, Mulrow CD, et al The PRISMA 2020 statement: an updated guideline for reporting systematic reviews. BMJ. 2021;372:71.

17. McGowan J, Straus S, Moher D, Langlois EV, O’Brien KK, Horsley T, et al Reporting scoping reviews-PRISMA ScR extension. J Clin Epidemiol. 2020;123:177–9.

18. Warkentien T, Crum-Cianflone NF. An update on Cryptococcus among HIV-infected patients. Int J STD AIDS. 2010;21(10):679–84.

19. Hallgren KA. Computing Inter-Rater Reliability for Observational Data: An Overview and Tutorial. Tutor Quant Methods Psychol. 2012;8(1):23–34.

20. Landis JR, Koch GG. The measurement of observer agreement for categorical data. Biometrics. 1977;33(1):159–74.

21. Hong QN, Fàbregues S, Bartlett G, Boardman F, Cargo M, Dagenais P, et al The Mixed Methods Appraisal Tool (MMAT) version 2018 for information professionals and researchers. Education for Information. 2018;34(4):285–91.

